# Meditation Experience is Associated with Increased Structural Integrity of the Pineal Gland and greater total Grey Matter maintenance

**DOI:** 10.1101/2024.03.04.24303649

**Authors:** Emanuele RG Plini, Michael C Melnychuk, Paul M Dockree

**Affiliations:** Department of Psychology, Trinity College Institute of Neuroscience, Trinity College Dublin, Llyod Building, 42A Pearse St, 8PVX+GJ Dublin, Ireland

## Abstract

Growing evidence demonstrates that meditation practice supports cognitive functions including attention and interoceptive processing, and is associated with structural changes across cortical networks including prefrontal regions, and the insula. However, the extent of subcortical morphometric changes linked to meditation practice is less appreciated. A noteworthy candidate is the Pineal Gland, a key producer of melatonin, which regulates circadian rhythms that augment sleep-wake patterns, and may also provide neuroprotective benefits to offset cognitive decline. Increased melatonin levels as well as increased fMRI BOLD signal in the Pineal Gland has been observed in mediators vs. controls. However, it is not known if long-term meditators exhibit structural change in the Pineal Gland linked to lifetime duration of practice. In the current study we performed Voxel-based morphometry (VBM) analysis to investigate: 1) whether long-term meditators (LTMs) (n=14) exhibited greater Pineal Gland integrity compared to a control group (n=969), 2) a potential association between the estimated lifetime hours of meditation (ELHOM) and Pineal Gland integrity, and 3) whether LTMs show greater Grey Matter (GM) maintenance (BrainPAD) that is associated with Pineal Gland integrity. The results revealed greater Pineal Gland integrity and lower BrainPAD scores (younger brain age) in LTMs compared to controls. Exploratory analysis revealed a positive association between ELHOM and greater signal intensity in the Pineal Gland but not with GM maintenance as measured by BrainPAD score. However, greater Pineal integrity and lower BrainPAD scores were correlated in LTMs. The potential mechanisms by which meditation influences Pineal Gland function, hormonal metabolism, and GM maintenance are discussed – in particular melatonin’s roles in sleep, immune response, inflammation modulation, and stem cell and neural regeneration.

## Introduction

The pineal is a highly vascularised, singular, unpaired gland, and its most well-known function is the synthesis and release of the hormone melatonin, which regulates the sleep-wake cycle as well as other bodily rhythms. Melatonin is also involved in regulating mood, and it possesses immune and neuro-protective functions (Lee, 2019), regulates neural stem cell production (Yu, 2017) and is one of the most powerful and available anti-oxidants in the human body, particularly in the brain (Hardeland, 2021). There is also a concurrent popular belief, largely deriving from an occult or mystical perspective, that the pineal possesses a pivotal spiritual function, that it is a gateway to higher states of consciousness, and that it can be “activated” through various spiritual practices such as meditation or psychedelics. It is considered to be part of the physiological representation of the “third eye” by many people, most likely because it possesses anatomical similarities and shares phylogenetic history with the eye itself.

Putting mystical interpretation of pineal function aside, an interesting study (Liou, 2007) has nevertheless reported that the pineal gland, along with the entire corpora quadragemina, had considerably higher activation during meditation practice compared to quiet rest. Increased melatonin levels in meditators have also been observed in several studies (Thambyraja, 2023; Tooley, 2000; Harinath, 2004). These findings raise the possibility that the increased melatonin observed with meditation may to some extent be a result of potential remodelling of the pineal gland following repeated activation.

Melatonin levels generally decrease as we age (Karasek, 2001), and there is significant recent interest in the relationship of this age-related decrease with accelerated cognitive decline and neurodegenerative processes. Findings have suggested that individuals with Alzheimer’s Disease (AD) exhibit irregularities of melatonin and pineal gland function (Song, 2019; Roy, 2022). Plasma concentration of melatonin is linearly correlated with volume of the pineal (Nolte, 2009; Mahlberg, 2006; Mahlberg 2008), suggesting pineal integrity could play a role. Given the sleep regulatory and brain maintenance roles of melatonin this seems plausible. The ability of melatonin supplementation to halt or slow the effects of age-related brain deterioration is also encouraging (Li, 2022; Vincent, 2018).

In the present study we examined the structural integrity of the Pineal using voxel based morphometry (VBM) in a cross-section of meditators with varied levels of expertise. We argued that if the Pineal is activated consistently during meditative practice, a reasonable consequence could be structural remodelling of the pineal, and that these structural changes, if present, should exhibit a relationship with the total number of hours spent in meditation. As a secondary aim we also investigated whether meditators exhibited greater grey matter maintenance in comparison to controls, as earlier research has suggested (Luders et al. 2016; Adluru et al. 2020).

## Methods

### Participants and secondary data

Data for the meditator group (14 individuals, 11 females, 3 males, ages 28-66) came from Hasenkamp lab’s prior work (Hasenkamp et al. 2012a & 2012b). Among the 14 subjects, six predominantly practiced Shamatha, five engaged in other Tibetan styles (compassion and tong-len), and three focused on Vipassana. Notably, all meditation styles employed or included breath-focus meditation. Healthy control data originated from various sources. 135 young subjects (96 males, 39 females, ages 20-35) were from the LEMON dataset (Babayan et al. 2019 – collected at Max Planck Institute for Human Cognitive and Brain Sciences). MRI data were downloaded from https://fcon_1000.projects.nitrc.org/indi/retro/MPI_LEMON/downloads/download_MRI.html. 395 adult/elderly controls (168 males, 227 females, ages 56-95) were part of ADNI 3 phase from IDA (https://ida.loni.usc.edu). 398 individuals (249 males, 149 females) from BASE-II (Bertram et al. 2014) were accessed via https://www.mpib-berlin.mpg.de/research/research-centers/lip/projects/aging/base-ii). Lastly, 41 older adults (15 males, 26 females, ages 60-72) were provided by University of Dallas Texas’ Center of Brain Health (Chapman et al. 2016). These datasets, totalling 969 healthy controls, were compared to 14 meditators.

### Neuroimaging processing and BrainPAD calculation

All the images 3T high-resolution T1-weighted MRIs in Nifti format of the 14 meditators and the 969 healthy controls were processed using Computational Anatomy Toolbox 12 – CAT12 (http://www.neuro.uni-jena.de/cat/) implemented in Statistical Parametric Mapping 12 – SPM12 (https://www.fil.ion.ucl.ac.uk/spm/). The processing was run following the default CAT12 settings, except for the voxel size that was settled at 1mm isotropic voxel size. At completion of the processing the images total intracranial volume (TIV) was calculated, and the image were modulated and spatially oriented in the Montreal Neurological Institute – MNI coordinates space. All the images had a quality score above 75% - CAT12 reports of quality assurance rating scale. Subsequently, the processed normalized images Grey Matter (GM) + White Matter (WM) were smoothed with a 2 mm^3^ FWHM kernel.

Brain Predicted Age Discrepancy (BrainPAD) is an objective measure of brain aging, which reflects age-related Grey Matter (GM) maintenance. Negative values imply younger brain, positive values signal accelerated GM degeneration. Zero means no age difference between chronological and brain GM age. BrainPAD computes age disparity using defined GM trajectory in healthy subjects and was calculated here using the method of Boyle and colleagues (Boyle et al. 2020). Further details are provided in the supplementary materials.

### Pineal Gland ROI definition

In order to isolate the Pineal Gland in the analyses a binary mask oriented in the MNI space was manually developed (voxel by voxel in FSL eyes – edit mode) (https://fsl.fmrib.ox.ac.uk/fsl/fslwiki/). By using as anatomical reference the average brain template available in CAT12 (derived from 555 healthy control subject – “Template_T1_IXI555_MNI152_GS” - http://brain-development.org/), and considering the Pineal anatomical localization described in literature (Haines 2004; Mai and Paxinos 2012), on the coronal plane, the Pineal area was isolated voxel by voxel (1 mm^3^ isotropic voxel size) proceeding slice by slice until the boundaries of the Corpora Quadrigemina and using as reference the Pineal and Supra-Pineal Recesses. The binary (1 mm^3^ isotropic voxel size) did not encroach the Splenium of the Corpus Callosum and the Thalamus. The binary ROI had the matrix size (FOV) of 181×217×181 (the same of the processed images outcoming from CAT12).

### Structural Analyses in CAT12 and JASP

In a first instance (**1**^st^ **branch**) a Voxel-based morphometry (VBM) analysis investigated whether Meditators (group A) had greater Pineal integrity than controls (group B) via a two-sample t-test (controlling for age, gender and TIV). The main hypothesis was tested in SPM contrast manager (1 −1, group A > group B) isolating the Pineal Gland using a binary mask by applying progressive statistical thresholds from P<0.05 to stricter (P<0.01; P<0.001; P<0.05 FWE corrected).

In a second instance (**2**^nd^ **branch**), exploratory analysis investigated meditation hours’ impact on Pineal integrity. In a multiple regression model, ELHOM was entered as independent variable while controlling for age, gender, TIV. As in the first model, the Pineal region was isolated with a binary mask and the positive relationship with ELHOM was tested thresholding for P<0.05 to stricter. In the same way, another multiple regression model investigated a direct relationship between the Pineal Gland and BrainPAD.

Lastly (**3**^rd^ **branch**), in order to test whether meditators have reduced GM degeneration compared to the controls, in JASP, an ANCOVA model was created treating BrainPAD as a dependent variable, group as independent factor and age, gender and TIV were entered as covaried. Then post-hoc Bonferroni correction was applied. This analysis was carried out using 834 controls rather than 969, because Bayaban et al. 2019 refused to provide the exact age of the 135 participants of the LEMON, and it was therefore not possible to calculate BrainPAD.

## Results

**Table 1a** and **Table 1b** summarise the key demographic variables and brain parameters used in subsequent analyses.

**Table la:**
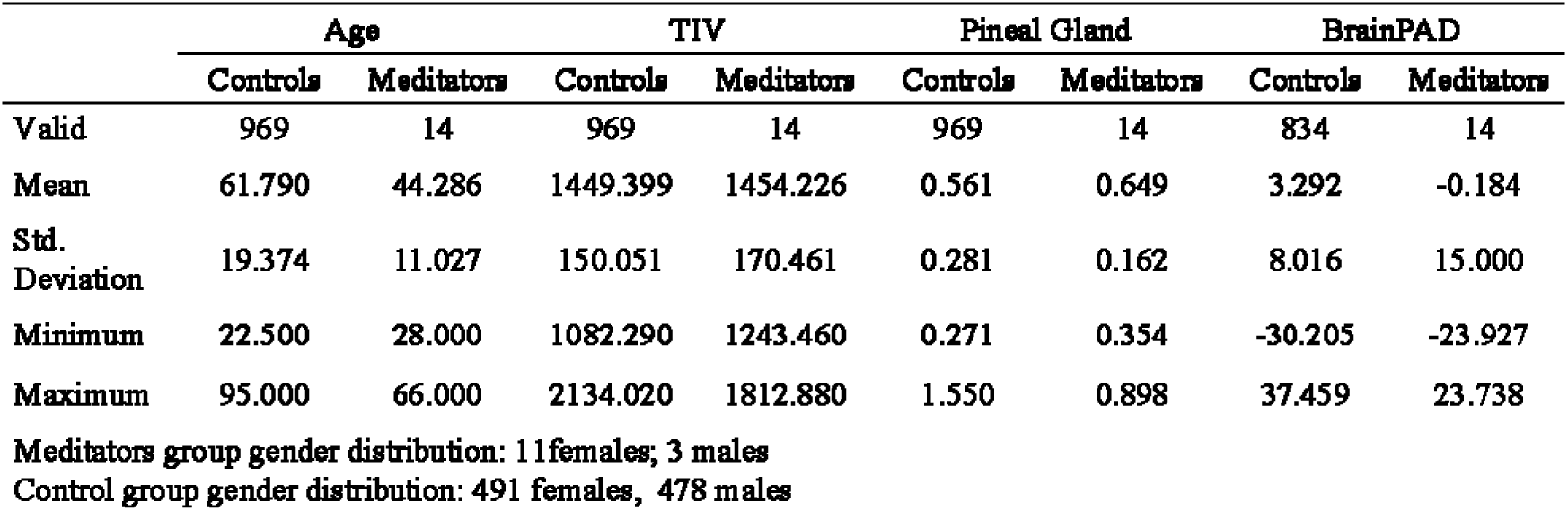
Groups descriptive Statistics.

**Table lb:**
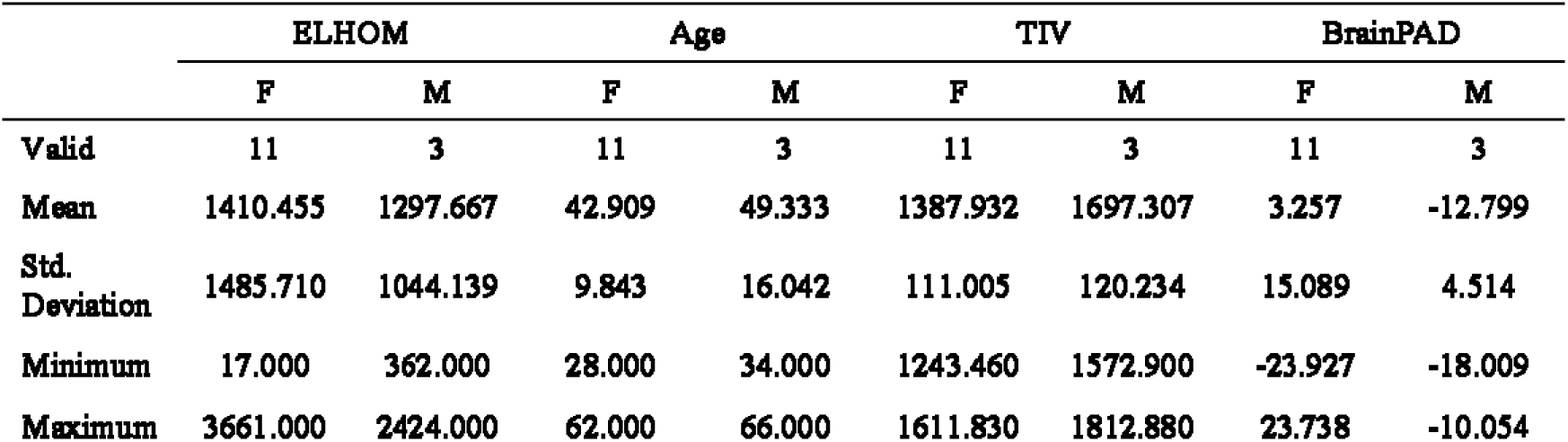
Meditators descriptive Statistics.

**Table 1a** reports mean Age, Total Intracranial Volume (TVA), Pineal Gland signal intensity and Brain Predicted Age Discrepancy, (BrainPAD) for the Meditation and Control groups. **Table 1b** reports the mean estimated lifetime hours of meditation (ELHOM), Age, Total Intracranial Volume (TVA) and Brain Predicted Age Discrepancy, (BrainPAD) stratified by sex.

### Differences between Pineal Gland Integrity in Meditators vs Controls

As reported in **Table 2**, by controlling for age, gender and TIV, in comparisons to 969 healthy controls, meditators group exhibited greater Pineal Gland signal intensity for the statistical threshold of P<0.05. In specific, a cluster of 363 voxels (T 5.40 – FWE corrected P<0.038) within the Pineal region was significantly associated with practice of meditation. As shown in **Figure 1** the significant cluster matched the spatial localization of the Pineal Gland in the MNI space. (original SPM12 outputs are available in the supplementary materials).

**Figure 1.**
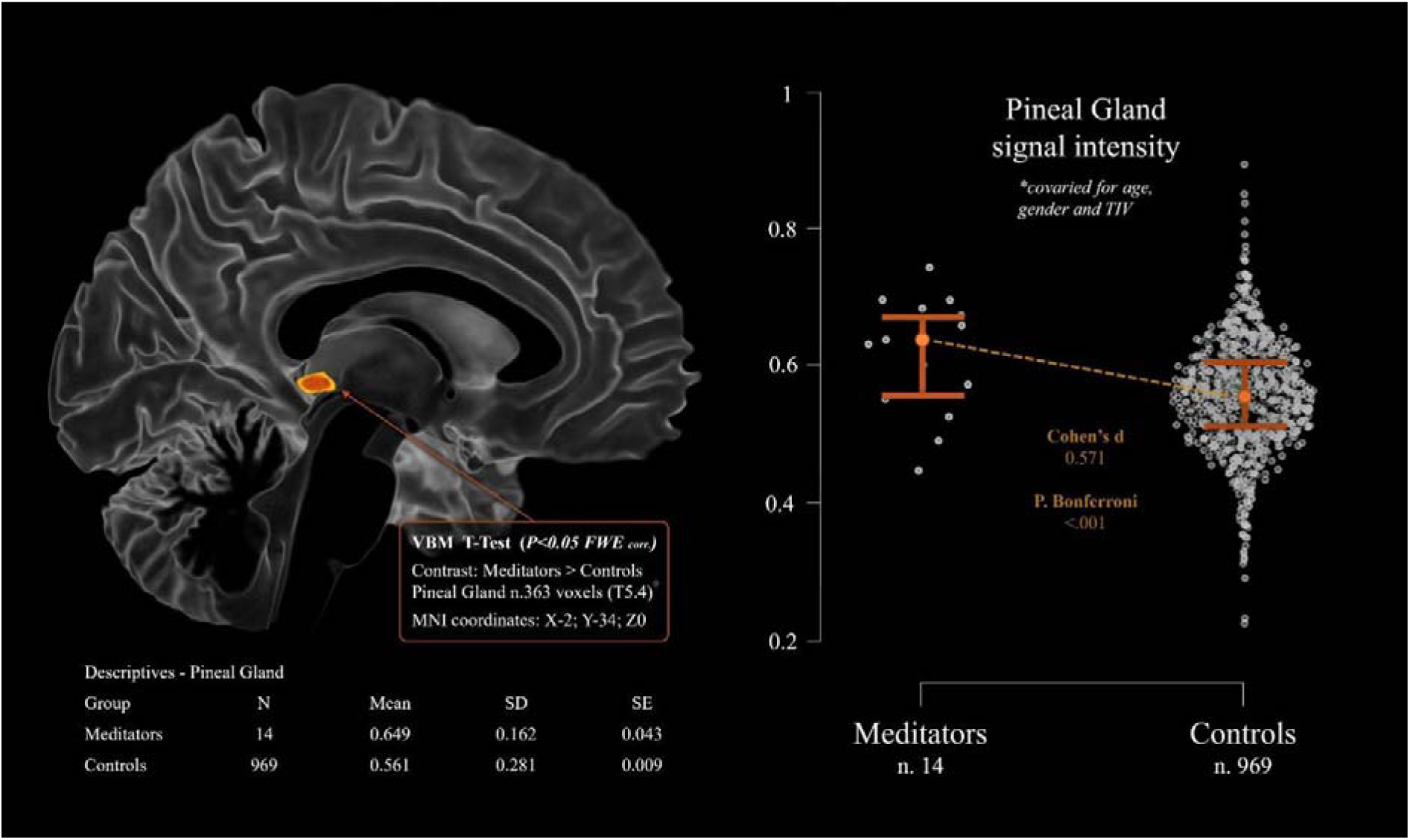
shows the significant cluster of voxels of the Pineal region where Meditator group exhibits greater signal intensity in comparison to the control group. The left portion shows the Pineal cluster (in yellow-orange) on axial 3D reconstruction of the brain. The right portion showcases the Pineal average signal intensity between the two groups. The model is corrected for age, gender and total intracranial volume.

#### MRI Structural analyses table: T Test

**Table 2.**
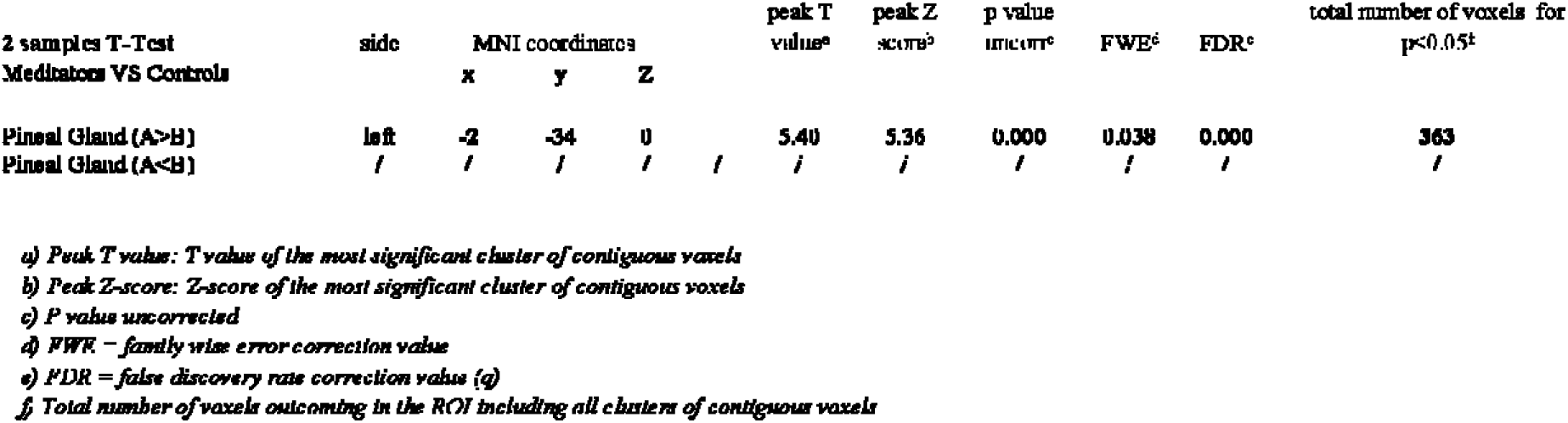
shows the results of the two-samples T-Test comparing the structural variation of the Pineal Gland between a group of 14 meditators (group A) and 969 healthy controls (group B). The analyses tabled both the hypothesis (Group A>B and Group B>A). The model was controlled for age, gender and total intracranial volume. The table reports the significant clusters of voxels for the differences observed between the two groups for the statistical threshold of P<0.05. Pineal Gland structural difference survived multiple comparison corrections when was applied (FWE corr. P<0.038)

### The Relationship between Pineal Gland signal intensity and hours of meditation

An exploratory analysis tested for an association between ELHOM and the Pineal Gland. For P<0.01 (uncorrected) a small cluster of 16 voxels (T = 3.33, BF_10_ = 1926.853, R² = 0.795) localised in the Pineal Gland was positively associated with more ELHOM, namely, the estimated hours of meditation practise was associated with greater signal intensity of the Pineal Gland (see Table 3 and Figure 2). Bayesian modelling suggests very strong evidence for this relationship (BF_10_ = 1926.853, R² 0.795). However, the cluster did not survive multiple comparison corrections (original SPM12 outputs are available in the supplementary materials). In addition, as reported in the supplementary materials, greater Pineal integrity was directly associated with lower BrainPAD scores (BF10 360.042 - R² 0.692).

**Figure 2.**
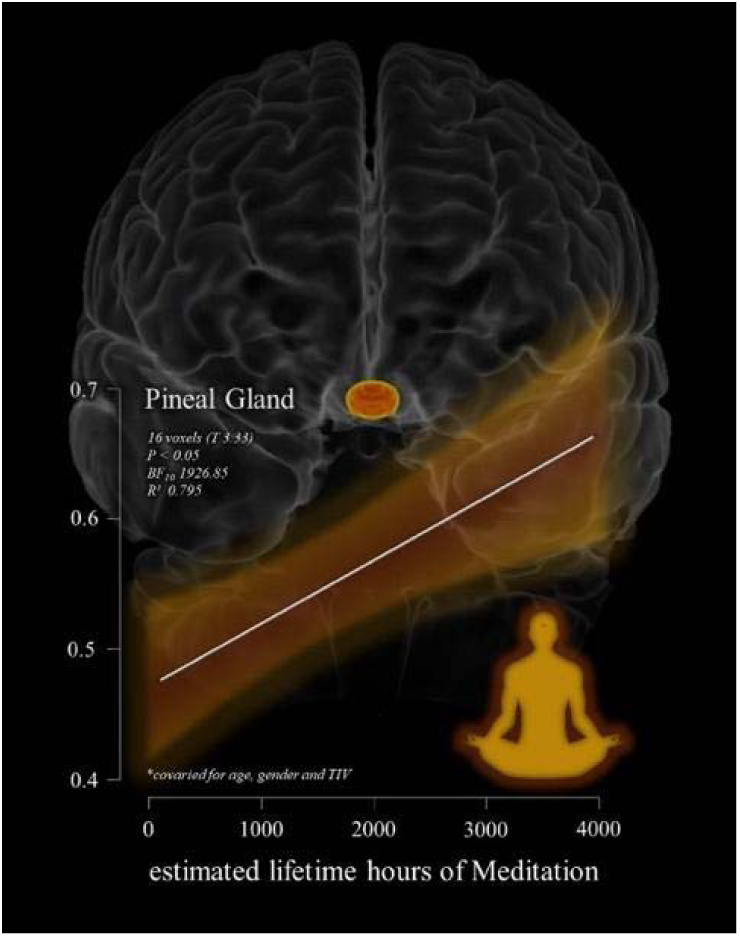
shows the relation between the significant cluster of voxels in the Pineal Gland (highlighted in yellow-orange on a 3D reconstruction of the brain – coronal view) associated with estimated lifetime hours of meditation (ELHOM) for the 14 individuals examined in the MRI structural multiple regression model. The model and the scatterplot are controlled for the effects of age, gender and total intracranial volume for the statistical threshold of P<0.05.

#### MRI Structural analyses table: Multiple Regression model - Pineal Gland and boors of meditation

**Table 3.** shows the results of the multiple regression model exploring potential positive between Pineal Gland structural single intensity and estimated lifetime hours of meditation (ELHOM) in a group of 11 meditators. The model was controlled for age, gender and total intracranial volume. The table report the significant cluster of voxels for the statistical threshold of P<0.05. Pineal Gland (and Supra-Pineal area) greater signal intensity is positive with greater number of hours of meditation. However, the result did not survive multiple comparison corrections when was applied.

| Multiple Regression<br>ELHOM | side | MNI coordinates | | | peak T<br>value<sup>a</sup> | peak Z<br>score<sup>b</sup> | p value<br>uncorr<sup>c</sup> | FWE<sup>d</sup> | FDR<sup>e</sup> | total number of voxels for $p < 0.05$<br>with max BF<sub>10</sub><sup>f</sup> |
| --- | --- | --- | --- | --- | --- | --- | --- | --- | --- | --- |
|  |  | x | y | z |  |  |  |  |  |  |
| Pineal Gland | right | 4 | -31 | 2 | 3.33 | 2.62 | 0.004 | 1.000 | 1.032 | 16 (BF<sub>10</sub> 1926.853 - R<sup>2</sup> 0.795) |

a) Peak T value: T value of the most significant cluster of contiguous voxels
b) Peak Z-score: Z-score of the most significant cluster of contiguous voxels
c) P value uncorrected
d) FWE – family wise error correction value
e) FDR – false discovery rate correction value (q)
f) Total number of voxels outcoming in the ROI including all clusters of contiguous voxels (in brackets are reported Bayes Factors)

### Differences in Grey Matter Maintenance in Meditators vs. Controls

An ANCOVA model, controlling for Age, Gender and TIV, demonstrated that Mediators (n=14) exhibited reduced GM degeneration in comparison with Controls (n=834). On average, Controls had a predicted brain age that was 3-years older than their chronological age, while meditators shown GM aging consistent with their chronological age (F = 14.13, p < .001 d = .97). In addition, as reported in the supplementary materials, greater Pineal integrity was directly associated with lower BrainPAD scores (BF_10_ = 360.042, R² 0.692). There was no association between BrainPAD scores and ELHOM.

**Table 4.**
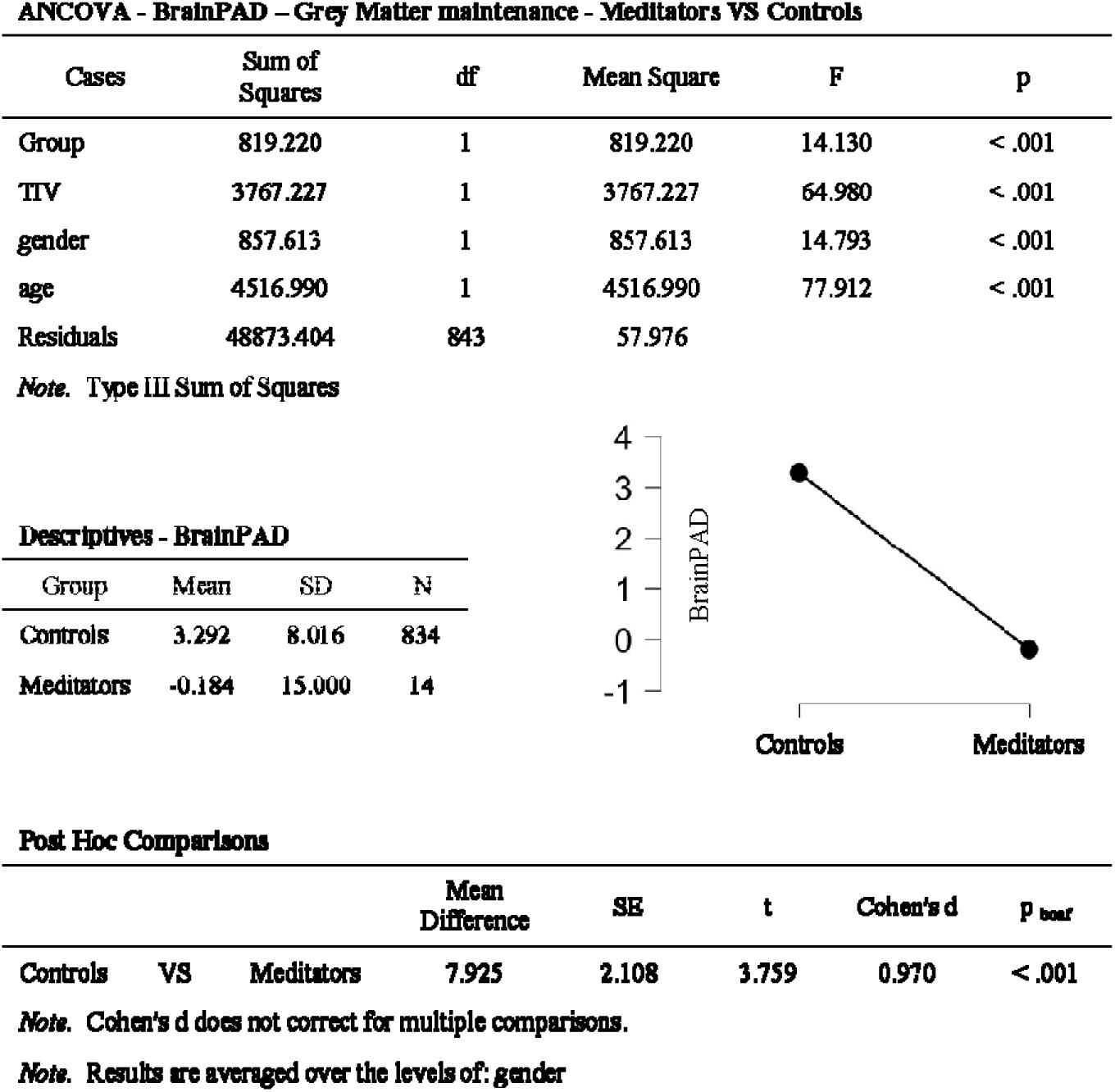
reporting the results of the ANCOVA model investigating differences in Grey Matter maintenance (BrainPAD) between Meditators VS Controls. Results shown that Meditators exihbited reduced GM deneration in comparison to controls, in average controls had a brain 3 years older than their chronological ąge, while meditators shown GM aging consistent with their chronological ąge.

**ANCOVA - BrainPAD – Grey Matter maintenance - Meditators VS Controls**
| Cases | Sum of Squares | df | Mean Square | F | p |
| --- | --- | --- | --- | --- | --- |
| Group | 819.220 | 1 | 819.220 | 14.130 | < .001 |
| TIV | 3767.227 | 1 | 3767.227 | 64.980 | < .001 |
| gender | 857.613 | 1 | 857.613 | 14.793 | < .001 |
| age | 4516.990 | 1 | 4516.990 | 77.912 | < .001 |
| Residuals | 48873.404 | 843 | 57.976 |  |  |

*Note.* Type III Sum of Squares

**Descriptives - BrainPAD**
| Group | Mean | SD | N |
| --- | --- | --- | --- |
| Controls | 3.292 | 8.016 | 834 |
| Meditators | -0.184 | 15.000 | 14 |

**Post Hoc Comparisons**
|  |  |  | Mean Difference | SE | t | Cohen's d | p<sup>bonf</sup> |
| --- | --- | --- | --- | --- | --- | --- | --- |
| Controls | VS | Meditators | 7.925 | 2.108 | 3.759 | 0.970 | < .001 |

*Note.* Cohen's d does not correct for multiple comparisons.
*Note.* Results are averaged over the levels of: gender

## Discussion

We have observed greater structural integrity of the pineal gland in meditators compared to controls, and this effect is linearly related to lifetime hours spent in meditation. We also observed better maintenance of GM (BrainPAD, a proxy measure indicating reduced “brain age”) in the meditation group relative to controls, in accordance with previous similar findings (Pagnoni & Cekic, 2007; Luders, et al., 2009; Vestergaard-Poulson, 2009; Luders, et al., 2015; Hernandez, 2020). Furthermore, we found that greater pineal gland signal intensity was associated with reduced brain age in meditators. While the evidence for pineal integrity is preliminary, it does suggest that meditation provides some structural augmentation of the pineal gland. Previous studies have shown an increased fMRI BOLD response in the pineal gland during meditation (Liou et al 2005) as well as higher plasma melatonin concentrations in advanced meditators vs. non-meditators (Solberg et al 2004), but the current findings are the first, to our knowledge, to demonstrate morphometric differences in this melatoninergic structure in long-term meditation practitioners. Our secondary findings, showing that grey matter maintenance is enhanced in meditators, is in keeping with meta-analysis showing preserved and/or increased grey matter across several cortical structures (Fox, 2014), (Marciniak 2014; Gard, 2014; Sumantry, 2021).

Although the mechanisms behind meditation practice and hormonal changes are poorly understood, there are several ways in which meditation might influence melatonin production and affect morphological changes in the pineal gland. First, meditation is frequently practiced in an undisturbed dimly lit environment and typically, although not exclusively, it involves closing the eyes. Reduced light exposure, particularly from artificial sources of light, may enhance melatonin production over time (Lewy et al 1980). However, a previous study has demonstrated greater plasma melatonin concentrations following meditation compared to an eyes-open control condition, where light exposure was below the minimum intensity known to influence melatonin levels (Tooley et al., 2000). The implication is that reduced light exposure alone is not sufficient for any meditation-related effect upon melatonin. Second, it is possible that sleep duration or quality is a third variable explaining the relationship between meditation and pineal signal intensity. The most well-known action of melatonin is its regulation of sleep and waking via its effects on circadian regulation. Both melatonin supplementation and meditation practice have both been shown to improve sleep quality (Nagendra, 2012), It is plausible that improved sleep patterns brought about by habitual engagement with meditation lead to more regular melatonin secretion, particularly higher night-time melatonin levels (Tooley et al, 2000), and subsequently greater pineal integrity, since melatonin plasma concentration is correlated linearly with pineal volume (Nolte, 2009; Mahlberg, 2006 and 2008; Liebrich 2013). Third, the stress-reducing or relaxation-promoting effects of meditation may also be important factors that facilitate melatonin release. It is known that high levels of anxiety can suppress melatonin production. Meditation practice may serve to counter this effect over time via anxiolytic action that stimulates greater melatonin production from the pineal gland. Fourth, respiration is a primary pneumatic driver of cerebrospinal fluid (CSF) dynamics (Dreha-Kulaczewski, et al., 2015), and yogic breathing practices are known to modulate CSF dynamics in a pulsatile fashion (Yildiz, et al., 2022). Beneficial changes in the regularity and depth of respiration resulting from meditation could have positive impacts on waste clearance in and around the pineal gland, which is directly exposed to the ventricular space. Similarly, increased CSF flow might augment distribution of melatonin which is released directly into the CSF. In addition, increased ventricular dynamics could generate a weak piezoelectric induction in the pineal via mechanical stimulation of calcite micro-crystals which are known to be present there (Lang, et al., 1996; Baconnier, et al., 2002). A final, possibility is that the habenular activation observed during meditation by (quadragemina study) induces activation of the pineal gland via a previously observed direct efferent pathway from the medial habenula to the pineal (Ronnekleiv & Moller, 1979; Herkenham & Nauta, 1979). Approximately 15% of pineal cells were indeed observed to respond to habenular activation in a murine model (Rønnekleiv, et al., 1980).

Irrespective of the potential mechanism, structural augmentation of the pineal gland and higher melatonin secretion over time in meditation practitioners is likely to have benefits for cognitive aging. Notably, our finding that greater pineal signal intensity is associated with a younger brain age suggests that greater brain maintenance over time might be underscored by the neuroprotective properties of melatonin secretion. As we age, the pineal is both reduced in size (Arunkumar, 2015) and produces less melatonin (Karasek & Reiter, 2001). Pineal volume and plasma melatonin concentration is further decreased in individuals with dementia (Matsuoka, 2018; Matsuoka, 2020, Liu, 1999; Skene, 2003). Below we briefly outline literature and findings concerning the pineal gland and melatonin that are most likely relevant to age-related neural degeneration and the concomitant cognitive and health benefits observed in meditators.

### Immune and Inflammatory Response Modulation

An array of immune and inflammatory processes are thought to play a role in early stages of neurodegeneration in Alzheimer’s Disease (AD). Melatonin is an immune modulator that displays both pro- and anti-inflammatory properties, through a complex host of different pathways. For example, pro-inflammatory actions of amyloid-β peptides are reduced by enhancing α-secretase and inhibition of β- and γ-secretase. In fact, melatonin is known to lower Aβ peptide levels (Lahiri, 1999, 2004; Garcia-Mesa, 2012;), and to inhibit amyloid plaque deposition in transgenic mouse models of AD. (Matsubara, 2003; Feng, 2004). Reductions of Aβ formation, which are coincident with an attenuation of neuroinflammation, have also been demonstrated in mouse models. However, the efficacy of treatment depend on administration during early onset, and clinical studies suggest that melatonin cannot reverse AD in humans (Hardeland, 2018) highlighting the need for earlier optimization of melatonin levels in older adults at risk of AD. In this regard, there is some evidence that melatonin can improve cognitive function in later-stage dementia (Roy, 2022). If optimal sleep patterns via melatonin levels and integrity could be maintained prior to or early in disease progression via meditation, this could have a positive effect on levels of cognitive reserve (Stern, 2002), and help delay or attenuate cognitive decline, through sleep maintenance, but also through other mechanisms of brain maintenance discussed here.

During sustained immune insults, microglia, part of the brain’s innate immune system, can become overactivated, leading to an excessive production of harmful inflammatory factors. Melatonin can regulate the glial response and reduce damage to neural cells resulting from oxidative stress and inflammation (Hardeland, 2021). All of these factors suggests that a lack of adequate and appropriate circulating melatonin can worsen pathological outcomes due to unregulated immune responses in the brain, and that maintaining these levels via a structurally preserved pineal gland could help delay or reduce the severity of neurodegeneration in pathological cases if addressed at an early stage.

### Anti-Oxidant Role

Reactive oxygen species are natural byproducts of cellular metabolism, and moderate levels are required for normal function, but an excess buildup of these reactive molecules damages cellular components. The resulting state of “oxidative stress” is associated with many disease states, including advanced ageing and neurodegeneration such as that seen in AD and Parkinson’s Disease (PD) (Butterfield, 2019). Antioxidants are a broad class of compounds and enzymes that neutralise excessive reactive oxygen molecules derived from metabolic processes and prevent ensuing tissue damage. Melatonin is an extremely powerful antioxidant (Tordjman, S., 2017; review: Lee, 2019), particularly in the brain, and decreased melatonin has been hypothesized as a contributing factor in tau deposition and AD pathogenesis (Wu, 2005). Melatonin supplementation has also been shown to inhibit and reduce tau pathology (Das, 2020). Minimising age-related decline in melatonin production via a meditation practice could have the potential to lower oxidative stress in the brain early-on in disease progression and affect the time-course of both age-related and pathological neural decline.

### Stem Cells, Neural Regeneration

Neural stem cells (NSCs), found in the anterior subventricular and subgranular zones of the hippocampal dentate gyrus, are capable of self-replication and differentiation into mature neurons, astrocytes, or oligodendrocytes, and contribute to neurogenesis even in the adult brain. Melatonin is a crucial regulator of NSC precursor cell commitment and differentiation (Chen, 2014), and it also promotes the proliferation of these NSCs in these areas (Moriya, 2007). Melatonin aids in NSC proliferation (via TrkB activation and the c-Raf-MEK-ERK1/2 pathway), and also facilitates survival of new neurons derived from precursor cells in the adult hippocampus (Ramirez-Rodriguez, 2009; Ortiz-Lopez, 2016). Melatonin regulates neural differentiation of induced pluripotent stem cells via melatonin receptor activation. Maintaining melatonin production and release at adequate levels is therefore desirable for neurogenesis and neural plasticity, and a regular meditation practice could assist in achieving this by both increasing baseline melatonin levels and maintaining or increasing pineal integrity.

### Limitations

The main limitations of the current study are the small sample size of the meditators and the cross-sectional nature of the models, which render our findings of preliminary interest but in need of further replication. Future studies should focus on gathering larger samples of meditators while performing longitudinal investigation together with neuropsychological testing, and sociodemographic information including diet and level of physical fitness. These multidimensional approach will help parse the role of meditation from other contribution factors to overall brain health and the related structural integrity.

Though we have replicated earlier evidence demonstrating that mediators show less grey matter degeneration, we observe a smaller statistical effect than reported by Luders et al. 2016 and Adluru et al. 2020. Nevertheless, it should be noted that this discrepancy might be attributable to the different methodologies behind the estimation of BrainAGE and BrainPAD in these studies. All studies report the same direction of effect in which meditative practice is associated with a younger and better maintained brain.

Although we demonstrate structural differences in the pineal gland dissociating meditators from controls and associating pineal gland signal intensity to hours of practice, it remains to be seen what specific elements of the contemplative experience are necessary for the observed morphometric effects. There are several variables that warrant further investigation including: diurnal light exposure, sleep duration, sleep quality and emotional health, all of which may contribute to structural modification of the pineal gland through melatonin secretion either directly from the processes of meditation or via collateral benefits of the practice.

The relationship between reduced GM degeneration and meditation may possibly mirror the effect of other relevant factors playing a role in overall brain health, such as the level of education, socio-economic status and a conceivable individual tendency of being more aware of healthy life-style choices (diet, physical fitness and meditation), which might be considered as alternative unconsidered dynamics underlying the neurophysiological effects of meditative experience. In the same vein, given the cross-sectional nature of the study, the possibility that morphological Pineal variations can induce to meditative practice should not be excluded, despite the fact we do not know of any potential mechanisms.

## Conclusions

Our current findings of greater pineal integrity, greater grey matter integrity, and lower brain age in meditators, combined with the large supporting body of literature on cortical/functional benefits of meditation, suggest that meditation could be beneficial in the context of cognitive and neurophysiological ageing by improving melatonin availability and regulation. Given melatonin’s downstream roles in sleep, free radical scavenging, anti-inflammation, immune modulation and neural stem cell proliferation and differentiation, it seems plausible that early pineal degeneration could play a causal role in age-related brain degeneration, and that structural preservation via meditation could be of benefit. More research is required, but the structural benefits to the pineal appear to be cumulative with practice, so how an acute intervention using meditation might be of benefit once cognitive and cortical degeneration have begun is not known. It seems likely that optimal benefit from mindfulness and breathing practices could be achieved from early and continuous practice across the lifetime.

## Author Contribution

**ERGP** : conceptualization, formal analyses, visualization, manuscript writing, interpretation. **M C M**: conceptualization, analyses support, manuscript writing, interpretation supervision. **PMD** : conceptualization, manuscript edits, supervision.

## Funding and Acknowledgments

Project founded by the Irish Research Council—Irish Research Council Laureate Consolidator Award (2018-23) IRCLA/2017/306 to Paul Dockree.

This work was further supported by ProApto which founded this study (more info: https://www.proapto-camouflage.com/)

This work reports data from the Berlin Aging Study II project, which was supported by the German Federal Ministry of Education and Research (Bundesministerium für Bildung und Forschung [BMBF]) under grant numbers #01UW0808, #16SV5536K, #16SV5537, #16SV5538, #16SV5837, #01GL1716A, and #01GL1716B. Another source of funding is the Max Planck Institute for Human Development, Berlin, Germany. Additional contributions (e.g., equipment, logistics, and personnel) are made from each of the other participating sites.

This reports data by the Centre of Brain Health – Dallas Texas which was supported by a grant from the National Institute of Health (RC1-AG035954, R01-NS067015, R01-AG033106) and by grants from the T. Boone Pickens Foundation, the Lyda Hill Foundation, and Dee Wyly Distinguished University Endowment.

Thanks are extended to Francesca Fabbricatore for proofreading the manuscript and for the thoughtful comments provided.

## Data availability statement

The Pineal Gland MNI mask developed for this study is available on reasonable request emailing Dr. Emanuele RG Plini ( –). The significant cluster of voxels of the Pineal Gland are available in nifti format in the supplementary materials.

The data on Meditators were provided by Wendy Hasenkamp as described in the methods and they are accessible upon reasonable request: https://wendyhasenkamp.net/

The ADNI and the LEMON datasets used are opensource and can be accessed referring to the information provided in the methods.

ADNI: https://adni.loni.usc.edu/data-samples/access-data/

LEMON: http://fcon_1000.projects.nitrc.org/indi/retro/MPI_LEMON.html

Data of the Berlin Aging Study II project can be accessed at the following link prior dedicated documentation: https://www.base2.mpg.de/7549/data-documentation

Data of the Dallas Centre of Brain Health: are available from Jeffrey S. Spence, Center for BrainHealth, University of Texas at Dallas, on reasonable request: https://centerforbrainhealth.org/

